# From HHV-6 reactivation to autoimmune reactivity against tight junctions and neuronal antigens, to inflammation, depression, and chronic fatigue syndrome due to Long COVID

**DOI:** 10.1101/2024.06.28.24309682

**Authors:** Michael Maes, Abbas F. Almulla, Xiaoou Tang, Kristina Stoyanova, Aristo Vojdani

## Abstract

**Background:** Inflammation and autoimmune responses contribute to the pathophysiology of Long COVID, and its affective and chronic fatigue syndrome (CFS) symptoms, labeled “the physio-affective phenome.”

**Objectives:** To investigate whether Long COVID and its physio-affective phenome are linked to autoimmunity to the tight junction proteins, zonulin and occludin (ZOOC), and immune reactivity to lipopolysaccharides (LPS), and whether the latter are associated with signs of human herpes virus-6 reactivation (HHV-6), autoimmunity directed against oligodendrocyte and neuronal proteins, including myelin basic protein (MBP).

**Methods:** IgA**/**IgM/IgG responses to Severe Acute Respiratory Syndrome Coronavirus 2 (SARS-CoV-2), HHV-6, ZOOC, and neuronal proteins, C-reactive protein (CRP) and advanced oxidation protein products (AOPP), were measured in 90 Long COVID patients and 90 healthy controls. The physio-affective phenome was conceptualized as a factor extracted from physical and affective symptom domains.

**Results:** Neural network identified IgA directed to LPS (IgA-LPS), IgG-ZOOC, IgG-LPS, and IgA-ZOOC as the most important variables associated with Long COVID diagnosis with an area under the ROC curve of 0.755. Partial Least Squares analysis showed that 40.9% of the variance in the physio-affective phenome was explained by CRP, IgA-MPB and IgG-MBP. A large part of the variances in both autoimmune responses to MBP (36.3-39.7%) was explained by autoimmunity (IgA and IgG) directed to ZOOC. The latter was strongly associated with indicants of HHV-6 reactivation, which in turn was associated with increased IgM-SARS-CoV-2.

**Conclusions:** Autoimmunity against components of the tight junctions and increased bacterial translocation may be involved in the pathophysiology of Long COVID’s physio-affective phenome.

## Introduction

According to the latest data, it has been found that there are at least 65 million people worldwide who are affected by Long COVID disease [1]. A considerable number of individuals who have recovered from COVID-19 have reported persistent neuro-psychiatric symptoms [1, 2]. Neurological disease, encompassing both the central and peripheral nervous systems, is present in over 33% of individuals with long COVID [2–5]. Neuropsychiatric symptoms include chronic fatigue syndrome (CFS), depression, and anxiety that can last for up to 12 months after recovery [2–4].

In separate publications authored by some of the current authors, a validated latent vector was extracted from various neuropsychiatric symptoms such as CFS, depression, and anxiety due to Long COVID [6–8]. This latent construct was given the name the “physio-affective phenome of Long COVID.” Decreased oxygen saturation (SpO2) and elevated peak body temperature (PBT) during the acute phase of illness can predict the physio-affective phenome of Long COVID [9]. Both PBT and lower SpO2 are indicators that can be used to assess the severity of the immune-inflammatory response during acute infection [10].

Nonetheless, there remains a significant ongoing discussion regarding the underlying causes of Long COVID and the extent of its impact on individuals’ physical and mental well-being, including symptoms such as CFS, depression, and anxiety. Some of the authors of the present study have discovered molecular pathways that are involved in the development of symptoms in individuals with Long COVID disease. These pathways include the activation of the immune-inflammatory system (IRS), specifically the NLRP3, M1 macrophage, T helper (Th)-1, and Th-17 activation [6–8, 11–13]. Additionally, some of these authors found activation of the compensatory immunoregulatory system, as well as oxidative and nitrosative stress reactions. Furthermore, there was an observed increase in insulin resistance, a decrease in tryptophan levels, and an increase in tryptophan catabolites, such as kynurenine [11, 12]. A recent meta-analysis has found that Long COVID disease is associated with elevated levels of C-reactive protein (CRP), D-dimers, lactate dehydrogenase, leukocytes, lymphocytes, and interleukin (IL)-6 [14].

Emerging research suggests a potential link between the prolonged presence of Severe Acute Respiratory Syndrome Coronavirus 2 (SARS-CoV-2) and the development of Long COVID in some individuals [15–17]. In addition, Vojdani and colleagues found that Long COVID, and its physio-affective phenome exhibit elevated levels of immunoglobulins (Ig) IgG and IgM responses directed towards the spike protein of SARS-CoV-2 and Human Herpesvirus type 6 (HHV-6). This is evident from the increased IgG/IgM responses to HHV-6 and deoxyuridine 5′-triphosphate nucleotidohydrolase (HHV-6-duTPase) in Long COVID [15]. Therefore, these authors concluded that reactivation of dormant viruses could potentially contribute to the development of Long-COVID. In their study, Almulla et al. [18] found that heightened autoimmune responses, specifically those mediated by IgG and IgM to neuronal antigens, play a significant role in the development of Long COVID and the emergence of new affective and CFS symptoms. Some of the proteins mentioned in the study are myelin basic protein (MBP), myelin oligodendrocyte glycoprotein (MOG), neurofilament light (NFL), synapsin, tubulin, cerebellar protein-2, and the blood-brain barrier (BBB) proteins claudin-5 and S100B [18].

Thus, it can be inferred that infection with the SARS-CoV-2 virus has the potential to reactivate dormant viruses. This, in turn, may lead to heightened oxidative and nitrosative stress, weakened antioxidant defenses, activation of the immune response system, and an elevated risk of microglial activation and neuroinflammation. These processes can result in damage to neuronal cells and an increased production of antibodies targeting neuronal antigens [15, 16, 18].

Major depression (MDD) and CFS exhibit heightened oxidative and nitrosative stress, diminished antioxidant defenses, activation of IRS, and damage to neuronal antigens [19–22]. There is evidence to suggest that both MDD and CFS are associated with symptoms of leaky gut or increased gut permeability, which can lead to bacterial translocation [23–26]. Both disorders show elevated levels of IgA and IgM antibodies to various Gram-negative gut commensal bacteria, indicating increased bacterial translocation due to leaky gut. Changes in additional biomarkers related to intestinal permeability, such as intestinal fatty acid binding protein (I-FABP), have been observed in individuals with MDD and suicidal tendencies [27]. Furthermore, it has been suggested by Maes et al. [28] that there is evidence pointing towards a strong link between increased bacterial translocation in individuals with MDD and heightened IgM- and IgG-mediated autoimmune responses. Additionally, Rudzki and Maes [29] have proposed a link between leaky gut and microglial activation in MDD.

Abnormalities in gut permeability can be linked to disruptions in the tight junctions of the paracellular pathways, particularly occludin [26]. Several factors can disrupt the healthy barrier function of the body. These include gut dysbiosis, the use of antibiotics and immunomodulatory drugs, autoimmune responses to barrier structures, and the presence of zonulin [23–25, 30]. Zonulin, also known as pre-haptoglobin-2, plays a role in the regulation of tight junctions and, therefore, immune tolerance [31]. However, excessive production of zonulin can lead to an increase in intestinal permeability, commonly referred to as leaky gut [26, 32]. It is worth mentioning that HHV-6 infection or reactivation has been linked to gastrointestinal symptoms and infection of the gastroduodenal mucosa, as observed in studies by Halme et al. [33] and Amo et al. [34]. Therefore, it is plausible that the reactivation of HHV-6 in individuals with COVID-19 could potentially lead to gastrointestinal infection, resulting in a compromised gut barrier and the translocation of bacteria. This, in turn, may trigger autoimmune reactions targeting neuronal antigens.

There is still much to be discovered about the potential connection between Long COVID and indicators of leaky gut or autoimmune reactions to occludin and zonulin. Additionally, it remains unclear whether the latter are linked to signs of SARS-CoV-2 persistence, HHV-6 reactivation, autoimmunity to neuronal antigens, and the physio-affective phenome of COVID-19. Therefore, the aim of this study is to investigate whether Long COVID and its physio-affective phenome are linked to heightened autoimmune reactions directed against zonulin and occludin, and whether indicants of tight junctions dysfunction is connected to the reactivation of HHV-6 and autoimmune responses targeting neuronal antigens such as MBP, MOG, cerebellar-protein-2, synapsin, tubulin, NFP, and BBB-brain damage (BBD) proteins (claudin-5 and S100B).

## Participants and Methods

### Participants

In the present investigation, patients were recruited based on the Long COVID disease criteria established by the World Health Organization [35] and assessed by expert clinicians. Patients with long COVID are defined as individuals who have had a confirmed COVID-19 infection (see below) and have experienced at least two of the specified symptoms for a minimum of two months after the initial infection. These symptoms include emotional distress, concentration difficulties, cognitive dysfunctions, chronic fatigue, loss of sense of smell or taste, chest discomfort, persistent cough, difficulty breathing, headache, dizziness, muscle and joint pain, sleep problems, and gastrointestinal symptoms (WHO). These symptoms may continue beyond the initial phase of SARS-CoV-2 infection or become noticeable 2-3 months after the initial infection. All patients had Long COVID symptoms that persisted for at least 3 months.

This study enrolled a total of 180 individuals, namely 90 individuals diagnosed with Long COVID and 90 individuals who served as controls. The study sample was used to examine the differences in biomarkers among subjects with the diagnosis of Long COVID in comparison to healthy controls. In addition, we examined the associations between the leaky gut biomarkers and either autoimmunity biomarkers or antibodies to viral antigens. Healthy control serum samples (n=76) were supplied by Innovative Research, Novi, Michigan, and Immunosciences Lab., Inc. (Los Angeles, California, USA) included 54 Long COVID patients. The Iraqi site included 14 heathy controls and 58 Long COVID patients. The diagnosis of the acute stage of COVID-19 infection was always made by expert virologists and doctors. All subjects had been diagnosed during the acute infectious phase using a positive reverse transcription real-time polymerase chain reaction (rRT-PCR) test and the detection of IgG/IgM antibodies against SARS-CoV-2. The SARS-CoV-2 IgG antibody Elisa kit (Zeus, Scientific) was used to confirm that all control samples tested negative.

In the 72 Iraqi individuals, we measured the effects of SpO2 (Oxygen Saturation) and PBT (peak body temperature) during the acute phase of COVID-19 infection, and different rating scales to assess the physio-affective phenome of Long COVID. The Iraqi Long COVID patients received medical treatment from multiple healthcare institutions, namely Imam Sajjad Hospital, Hassan Halos Al-Hatmy Hospital for Infectious Diseases, Middle Euphrates Oncology Center, Al-Najaf Educational Hospital, and Al-Sader Medical City, all situated in Najaf, Iraq. We omitted all participants who had previously experienced chronic fatigue syndrome (CFS), major depressive episodes, dysthymia, bipolar disorder, schizophrenia, schizo-affective disorder, psycho-organic syndromes, panic disorder, generalized anxiety disorder, and substance use disorders (excluding nicotine dependence). We excluded all subjects (patients and controls) with established neuro-immune disorders such as Alzheimer’s disease, multiple sclerosis, stroke, and Parkinson’s disease, and systemic immune disorders such as psoriasis, chronic obstructive pulmonary disease (COPD), scleroderma, chronic kidney disease, and psoriasis. We also excluded pregnant or lactating women.

The Ethics Committee of the College of Medical Technology at the Islamic University of Najaf in Iraq has granted authority to conduct this research (Document No: 34/2023). Prior to being included in the study, all participants or their legal representatives gave written informed consent. The study’s design and implementation followed the standards set by the International Conference on Harmonization of Good Clinical Practice, the Belmont Report, the Council of International Organizations of Medicine (CIOMS) Guideline, as well as ethical and privacy rules in Iraq and globally. Our institution’s institutional review board adheres to the International Guidelines for the Conduct of Safe Human Research (ICH-GCP).

### Clinical measurements

In the Iraqi individuals, a paramedical specialist recorded PBT and SpO2 during the acute infectious phase using a digital sublingual thermometer with an audible signal and an electronic oximeter, both manufactured by Shenzhen Jumper Medical Equipment Co. Ltd. An expert psychiatrist conducted interviews with all participants approximately 3-4 months after the participants had recovered from the acute phase of SARS-CoV-2 infection. Data, including socio-demographic and clinical features, were obtained in this interview from individuals who participated in this study. The severity of the Long COVID phenome was determined by calculating a principal component score extracted from 4 different affective and CFS symptom rating scales. The severity of anxiety symptoms was assessed using the Hamilton Anxiety Rating Scale (HAMA) [36]. The severity of depressive symptoms was assessed using two well-established measures: the Hamilton Depression Rating Scale [37] and the Beck Depression Inventory-II (BDI) [38]. A psychiatrist utilized the Fibro-Fatigue scale to evaluate the extent of CSF [39].

The calculation of the body mass index (BMI) involves dividing an individual’s weight (in kilograms) by the square of their height (in meters). The diagnostic criteria for Tobacco Use Disorder (TUD) were sourced from the Diagnostic and Statistical Manual of Mental Disorders, Fifth Edition (DSM-5).

### Assays

Blood samples were drawn from fasting participants between 7:30 and 9:00 a.m. by venipuncture using vacutainer. The tubes were kept at room temperature for 15 minutes and then were centrifuged for 10 minutes at 2,000 RPM. The serum was separated into several smaller tubes and used in different assays.

Zonulin, occludin, alpha- and beta-tubulin were purchased from Abcam (Cambridge, MA, United States). Lipopolysaccharides (LPS) from *E. coli, Salmonella* and *Klebsiella*, and myelin basic protein (MBP) were purchased from Sigma-Aldrich (St. Louis, MO, USA). Neurofilament protein (NFP) was obtained from Bio-Techne R&D Systems (Minneapolis, MN, USA). Cerebellar protein-2 was purchased from CUSABIO (Houston, TX, USA). Myelin oligodendrocyte glycoprotein (MOG) peptide 74-96 and claudin-5 were synthesized by Biosynthesis (Lewisville, TX, USA). S100B was purchased from Antibodies (Limerick, PA, USA).

Zonulin and occludin at a concentration of 100 μg/mL were prepared in 0.01 M phosphate buffer saline with a pH of 7.4. These proteins were diluted 1:100 in 0.1 M carbonate buffer, pH 9.5, and 100 µL were added to wells of microtiter plates and incubated for 12 hours at room temperature followed by incubation at 4 [ for 16 h. The plates were washed three times with 200 µL of Trisbuffered saline (TBS) containing 0.05% Tween 20 (pH 7.4). The non-specific binding of immunoglobulins was prevented by adding 200 µL of 2% bovine serum albumin (BSA) in TBS and incubated overnight at 4 [. Plates were washed as previously described and then serum samples (diluted 1:50 for IgA, 1:100 for IgG) in 1% BSA in TBS containing 0.05% Tween 20 (pH 7.4) were added to duplicate wells and incubated for 1 h at room temperature. Plates were washed 5 times with 0.01 M PBS buffer to remove non-specific binding of serum components to the antigens. We then added 100 microliters of secondary antibodies labeled with alkaline phosphatase to each well. These antibodies (from Jackson ImmunoResearch, West Grove, PA, USA) were affinity-purified anti-human IgG FCy-specific, anti-human IgM FCμ-specific, and anti-human IgA alpha chain-specific. In this assay, the dilution in serum diluent buffer for anti-human IgG was 1:800, for anti-human IgM was 1:600, and for anti-human IgA was 1:200. After incubation, washing and addition of substrate, the color development was stopped by adding 50 microliters of 1 N NaOH. The intensity of the color was measured using an ELISA reader at a wavelength of 405 nm, and the indices were calculated using calibrators and control sera. Similarly, antibodies against SARS-CoV-2 superantigen, HHV-6 U24, and HHV-6 dUTPase, MBP, MOG, NFP, synapsin, cerebellar protein-2, and tubulin were measured using enzyme-linked immunosorbent assay (ELISA) as explained previously [15, 18].

The advanced oxidation protein products (AOPP) in serum were quantified using enzyme-linked immunosorbent assay (ELISA) kits from Nanjing Pars Biochem Co., Ltd. in Nanjing, China. The CRP latex slide test, a product manufactured by Spinreact® in Barcelona, Spain, was utilized to conduct measurements of CRP in human serum.

### Statistical analysis

The Pearson correlation coefficients were employed to examine the associations between IgA/IgM/IgG responses to diverse antigens and neuropsychiatric symptom scores.

The present study employed an analysis of variance (ANOVA) to compare continuous variables across different study groups, whereas a contingency table analysis was used to examine categorical variables. In order to identify the main factors that contribute to CFS and emotional symptoms in individuals with Long COVID, we performed multivariate regression analysis, considering age, gender, and BMI as controlling variables. A manual and an automated stepwise approach was used; a p-value of 0.05 was used as the criteria for including variables, while a p-value of 0.10 was used as the criteria for excluding variables. The model’s key metrics, including F, df, and p-values, as well as the total variance (R^2^) and standardized beta coefficients, were computed. We assessed the variance inflation factor (VIF) and tolerance in order to resolve any potential problems related to collinearity. In order to assess heteroskedasticity, we employed the White and modified Breusch-Pagan tests. In our study, we utilized binary logistic regression analysis to evaluate the associations between IgA/IgG/IgM responses and the diagnosis of Long COVID. We used healthy controls as the reference category. These analyses have accounted for potential confounding factors, such as age, sex, and study site. We calculated the standard error (SE), Wald statistics with p-values, the Odds ratio with 95% confidence intervals (CI), the classification table, and the Nagelkerke pseudo-R square (used to estimate the effect size). All tests were performed with a significance threshold of 0.05, using a two-tailed approach.

Neural networks were used to predict the diagnosis of Long COVID or the IGA/IgM/IgG responses to MBP or ZOOC as output variables and with other biomarkers serving as input variables. An automatic feedforward network model was used which consisted of either one or two hidden layers. The termination criterion was determined by observing a series of steps where the error term did not decrease any further. The models underwent training utilizing a maximum of 150 epochs in a batch-style training session. In case the output variable were two groups, we assessed the model’s predictive value by calculation the error, relative error, and the proportion of misclassifications. In case the output variable was a continuous score, we assessed the models’ predictive accuracy by comparing projected values to observed values using the coefficient of determination (R^2^) or the derived correlation coefficient. A significance chart was utilized to highlight the importance and relative significance of the input variables in the neural network model.

Principal Component Analysis (PCA) was employed to achieve feature reduction. An essential aspect of validating a principal component (PC) is to ensure that the explained variance (EV) reaches or exceeds 50%. In addition, it is important to ensure that the anti-image correlation matrix meets the necessary criteria. The factorability indicators should also be carefully evaluated, with a Kaiser-Meyer-Olkin (KMO) value that exceeds 0.65. It is crucial for Bartlett’s test of sphericity to yield a significant result, indicating that the variables are sufficiently correlated. Furthermore, all principal component loadings should surpass the threshold of 0.666. The statistical tests were performed using IBM’s SPSS application version 29, the most recent version available for Windows.

A comprehensive analysis was conducted using Partial Least Squares (PLS) to investigate the causal relationships between SARS-CoV-2 infection, HHV-6 reactivation, autoimmunity to ZOOC and neuronal antigens, and the phenome of Long COVID. The output variable (the physio-affective phenome) was derived from the clinical rating scale scores by extracting the first factor. A comprehensive PLS analysis is performed exclusively under certain circumstances: a) the factor extracted demonstrates satisfactory construct and convergence validity, as evidenced by a composite reliability of over 0.8, Cronbach’s alpha exceeding 0.7, and an average variance extracted (AVE) of more than 0.5; b) all loadings on the latent vectors are greater than 0.65 at a significance level of p < 0.001; c) the model exhibits a satisfactory fit, as indicated by the SRMR values being less than 0.08; d) Confirmatory Tetrad Analysis (CTA) indicates that the factor constructed is appropriately characterized as a reflective model. Through the utilization of 5,000 bootstraps, we computed path coefficients along with their corresponding p-values, specific indirect effects, and total effects.

Two power analyses were conducted using G*Power 3.1.9.7 to determine the minimum a priori sample size for detecting variations in logistic regression analysis (chi-square test; study part 1) and multiple regression analysis (study part 2). For study part 1, the power analysis showed that a minimum of 88 subjects is required. This calculation is based on an effect size of 0.3, a significance level (p) of 0.05, a power of 0.8, and degrees of freedom (df) equal to 1. For study part 2, the power analysis showed that a minimum of 65 subjects is required when considering an effect size of 0.25 (equivalent to 20% explained variance), a significance level of 0.05, a power of 0.8, and up to 7 predictors.

## Results

### Socio-demographic and clinical characteristics of Long COVID

Electronic Supplementary File Table 1 shows the sociodemographic and clinical data of the controls and Long COVID patients. There were no significant differences in age, sex, education, marital state, BMI, and smoking between both study groups. SpO2 was significantly lower in Long COVID as compared with controls, whilst PBT was significantly higher. All rating scale scores, including their composite score, were significantly higher in patients than in controls. The same table also shows that CRP and AOPP levels were significantly higher in patients than controls.

**Table 1.** Results of binary logistic regression analysis with the diagnosis Long COVID as dependent variable (heathy controls as reference group).

| Regression Number | Explanatory Variables | B | SE | Wald | P | OR | 95% CI |
| --- | --- | --- | --- | --- | --- | --- | --- |
| 1 | IgA LPS | 0.209 | 0.155 | 1.81 | 0.179 | 1.23 | 0.91; 1.67 |
| 2 | IgM LPS | -0.413 | 0.255 | 2.63 | 0.105 | 0.66 | 0.40; 1.09 |
| 3 | IgG LPS | 0.018 | 0.213 | 0.01 | 0.932 | 0.98 | 0.65; 1.49 |
| 4 | IgA ZOOC | 0.711 | 0.251 | 8.05 | 0.005 | 2.04 | 1.25; 3.33 |
| 5 | IgM ZOOC | 0.649 | 0.234 | 7.70 | 0.006 | 1.91 | 1.21; 3.03 |
| 6 | IgG ZOOC | 1.678 | 0.392 | 18.37 | <0.001 | 5.35 | 2.49; 11.53 |
| **BEST PREDICTION USING ALL IgA/IgG/IgM responses to LPS and ZOOC** |  |  |  |  |  |  |  |
| 7 | IgA ZOOC | 0.430 | 0.218 | 3.90 | 0.048 | 1.54 | 1.01; 2.36 |
|  | IgG ZOOC | 1.603 | 0.390 | 16.90 | <0.001 | 4.97 | 2.31; 10.66 |

OR: Odd's ratio, 95 CI: 95% confidence intervals, Ig: Immunoglobulin; LPS: lipopolysaccharides; ZOOC: zonulin and occludin.

### Ig responses to LPS and ZOOC in Long COVID

**Table 1** shows the results of binary logistic regression analysis that examined the associations between IgA/IgM/IgG responses to LPS and ZOOC and Long COVID. There were no significant associations between any of the Ig responses to LPS and Long COVID. All three Ig responses to ZOOC were significantly and positively associated with Long COVID. This table also shows the results of an automatic binary regression analysis that examined the effects of all six biomarkers shown in Table 1 on Long COVID. We found that Long COVID is predicted by the combined effects of IgG and IgA directed at ZOOC (Nagelkerke is 0.127).

Neural network analysis was employed to investigate the differentiation between Long COVID, and control groups based on the immune responses to LPS and ZOOC, both with and without immune responses to viral or neuronal antigens. The outcomes of the neural network analysis are displayed in **Table 2**. NN#1 utilizes the 6 Ig LPS and ZOOC data as input variables to differentiate between individuals with Long COVID and those without the condition. NN#1 selected the hyperbolic tangent as the activation function for the hidden layers and the identity function for the output layer. The training process involved two hidden levels, with the first layer consisting of 4 units and the second layer consisting of 3 units. Throughout the training process, the sum of squares error term was decreased, indicating improved ability of the neural network model to generalize trends. Based on the relatively constant error terms observed in the training, testing, and holdout samples, it can be concluded that the model has successfully reduced overfitting. The model’s precision, as determined through cross-validation was 66.7% with an AUC ROC curve 0.755. **Figure 1** illustrates the significance and standardized significance of all six input variables. The model primarily identified IgA-LPS, IgG-ZOOC, IgG LPS, and IgA ZOOC as the input variables with the most significant predictive capabilities. In contrast, IgM LPS and IgM ZOOC had a lesser impact on the model’s predictions.

**Figure 1.**
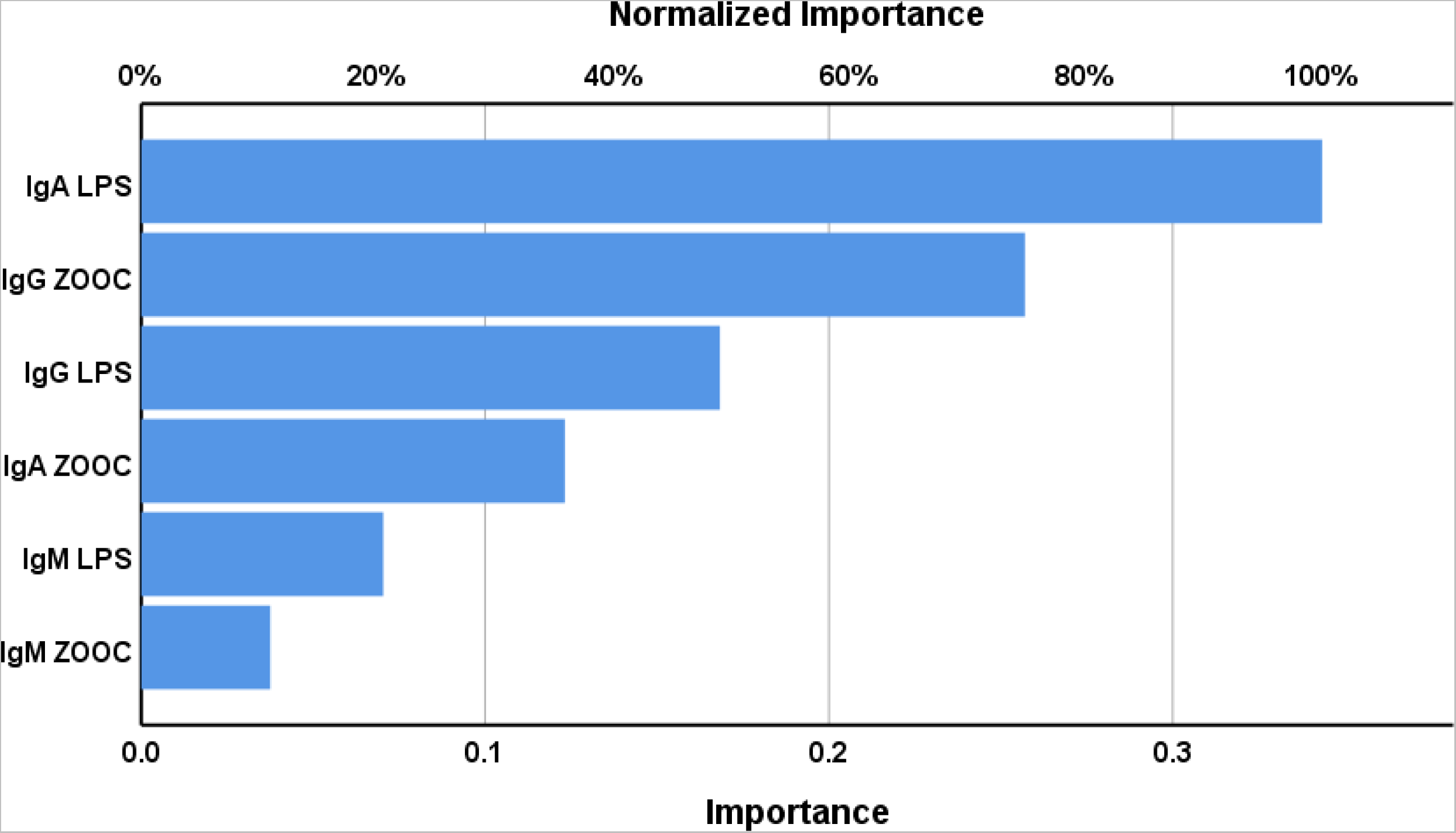
Results of neural network analysis showing the importance chart. The output variable is the diagnosis of Long COVID. Input variables are LPS: lipopolysaccharides; ZOOC: zonulin and occludin.

**Table 2.** Results of neural networks (NN) with Long COVID disease and healthy controls as output variables and IgA/IgM/IgG responses to lipopolysaccharides (LPS), zonulin and occludin (ZOOC) with or without other Ig responses to viral and neuronal antigens as input variables.

|  | Network information | NN#1 | NN#2 | NN#3 |
| --- | --- | --- | --- | --- |
| **Hidden layers** | Number of hidden layers | 2 | 2 | 2 |
|  | Activation function | Hyperbolic tangent | Hyperbolic tangent | Hyperbolic tangent |
|  | Number of units in hidden layer 1 | 4 | 6 | 6 |
|  | Number of units in hidden layer 2 | 3 | 5 | 5 |
| **Input variables** | Explanatory variables | IgA/IgM/IgG to LPS and ZOOC | IgA/IgM/IgG to LPS, ZOOC and viral antigens | IgA/IgM/IgG to LPS, ZOOC, viral and neuronal antigens |
| **Output variables** | Dependent variables | LC versus controls | LC versus controls | LC versus controls |
| **Output layer** | Number of units | 2 | 2 | 2 |
|  | Activation function | Identity | Identity | Identity |
| **Training** | Error term (sum of squares) | 17.933 | 18.634 | 12.539 |
|  | Percent incorrect | 29.5% | 24.6% | 12.5% |
| **Testing** | Sum of Squares error | 8.014 | 10.498 | 7.532 |
|  | Percent incorrect | 31.7% | 22.6% | 17.9% |
| **Holdout** | Percent incorrect | 33.3% | 22.6% | 10.2% |
| **ROC curve** | AUC | 0.755 | 0.863 | 0.947 |

Table 2, NN#2 utilizes the Ig LPS and ZOOC data, along with the viral antigens, as input variables to distinguish between Long COVID and control cases. Similar to NN#1, this neural network utilized the hyperbolic tangent as the activation function in the hidden layer and the identity function in the output layer. The training process involved the utilization of two hidden layers. The first layer consisted of 6 units, while the second layer contained 5 units. Based on the consistent relative error terms observed in the three samples (training, testing, and holdout), it can be concluded that the model does not exhibit signs of overfitting. Throughout the training process, there was a noticeable decrease in the sum of squares error term, indicating that the neural network model was able to generalize trends. The precision of the model, as determined through cross validation, was 77.4% (accuracy of the holdout sample). Additionally, the AUC ROC curve was found to be 0.863. Based on the data presented in **Figure 2**, it is evident that immune responses to HHV-6, ZOOC and LPS play a crucial role in predicting the occurrence of Long COVID compared to the control group. Following closely behind are IgM antibodies targeting ZOOC and HHV-6, IgA to SARS-CoC-2 and HHV-6 dUTPase, and LPS, and IgG to HHV-6 dUTPase and ZOOC. As such, the most important predictors of Long COVID are the leaky gut data coupled with immune responses to viral antigens, mostly HHV-6.

**Figure 2.**
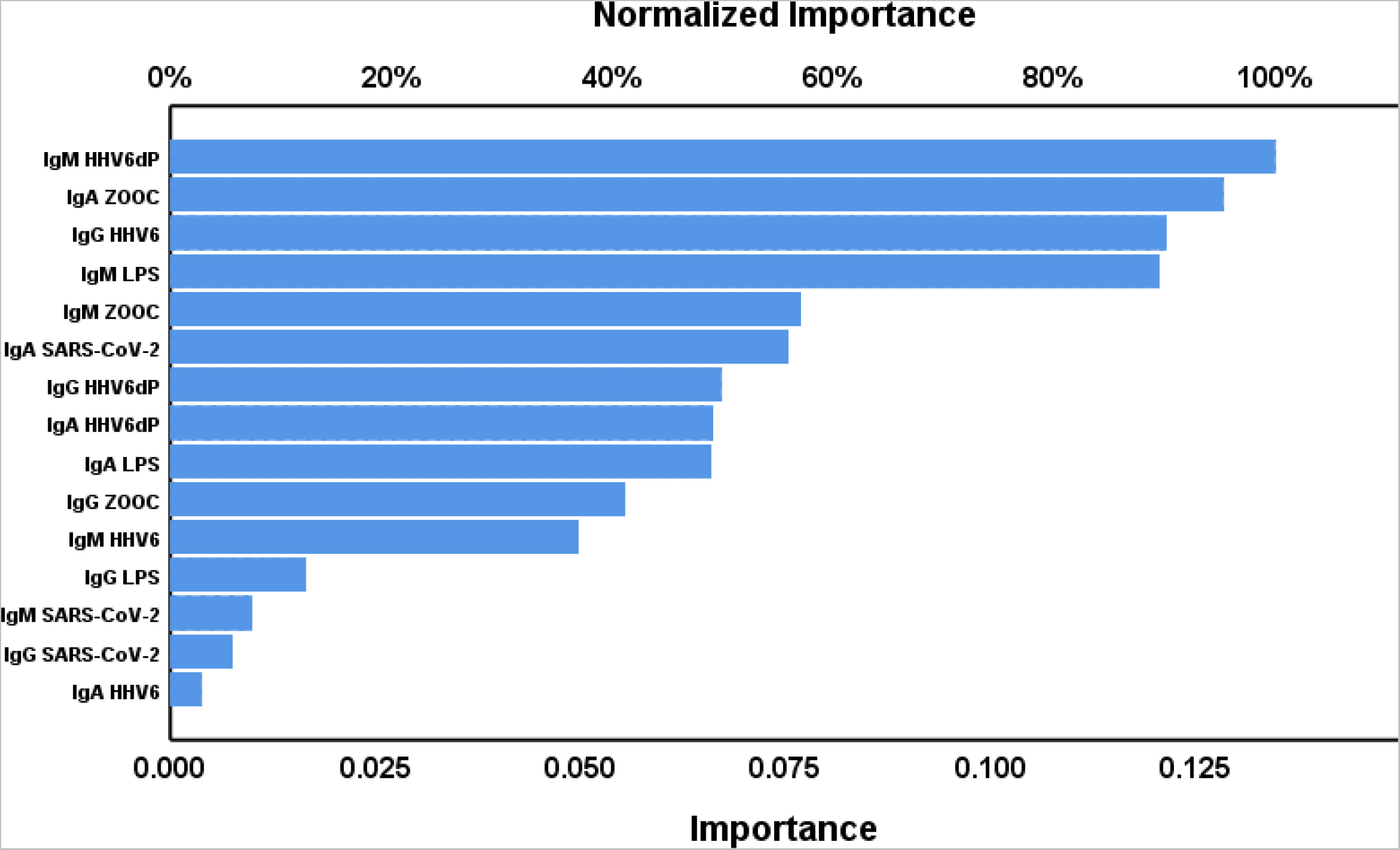
Results of neural network analysis showing the importance chart. The output variable is the diagnosis of Long COVID. Input variables are HHV6: Human Herpes Virus 6, HHV6dP: HHV-6 duTPase; LPS: lipopolysaccharides; ZOOC: zonulin and occludin; SARS-CoV-2: Severe Acute Respiratory Syndrome Coronavirus 2.

Table 2, NN#3 utilizes various immune responses as input variables, including those related to LPS, ZOOC, viral and neuronal antigens. The model was developed by reducing the number of input variables based on their relevance, as determined by the importance chart. Table 3, NN#3 presents all comprehensive network information, summary data, and confusion matrices. The results of the importance analysis are displayed in **Figure 3**. The precision of the model, when cross validated, surpassed that of NN#1 and NN#2. Specifically, it achieved an impressive 89.8% with an AUC ROC curve of 0.947. The most significant predictors of the model were IgG targeting HHV-6, BBB, MOG, synapsin, and ZOOC, IgA targeting BBB and ZOOC, and IgM targeting MBP, and HHV-6 dUPase.

**Figure 3.**
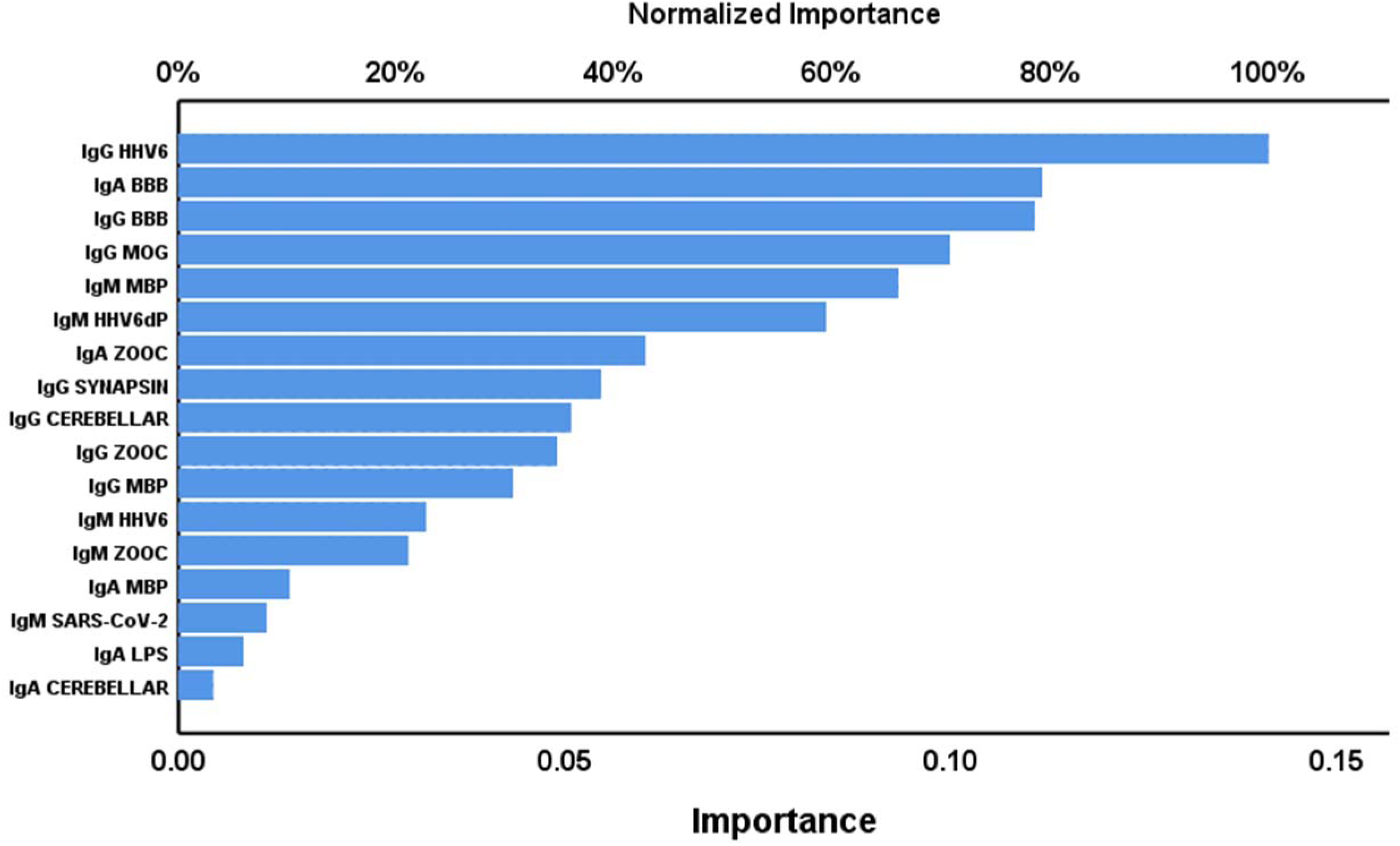
Results of neural network analysis showing the importance chart. The output variable is the diagnosis of Long COVID. Input variables are HHV6: Human Herpes Virus 6, HHV6dP: HHV-6 duTPase; BBB: blood brain barrier proteins; MOG: myelin oligodendrocyte glycoprotein; ZOOC: zonulin and occludin; cerebellar: Cerebellar protein-2; MBP: myelin basic protein; SARS-CoV-2: Severe Acute Respiratory Syndrome Coronavirus 2; LPS: lipopolysaccharides.

**Table 3.**
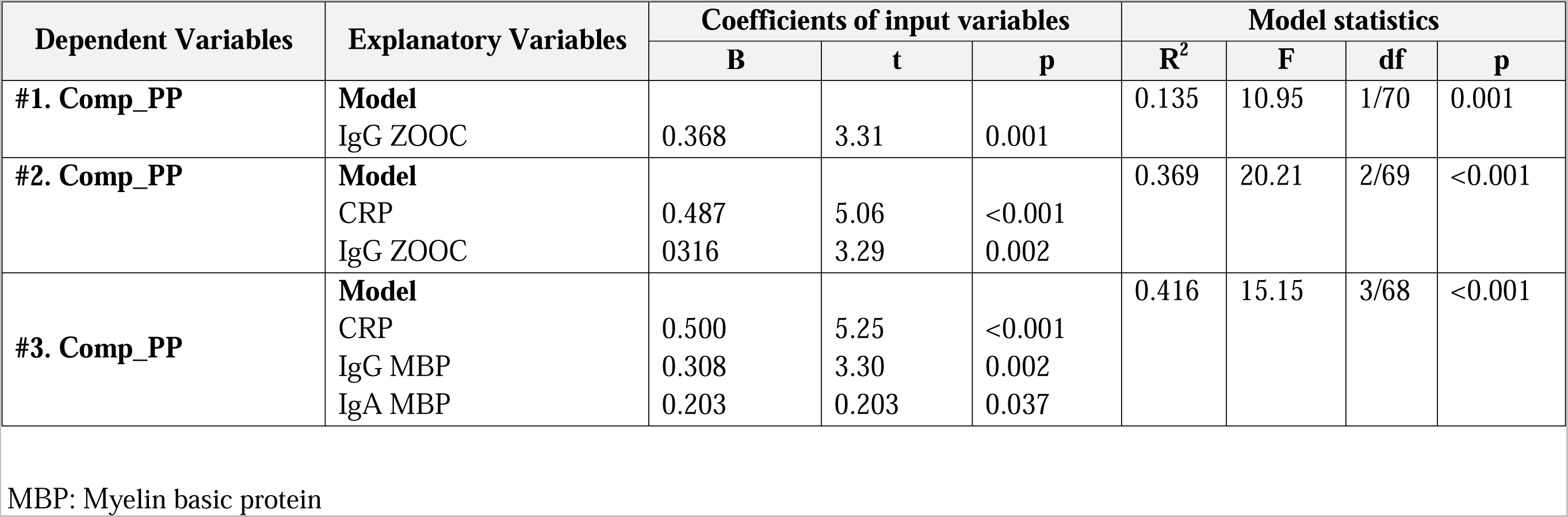
Results of multiple regression analysis with a composite phenome (Comp_PP) score as dependent variable and immunoglobulins (Ig) directed against lipopolysaccharides (LPS), zonulin and occludin (ZOOC), and neuronal proteins, levels of C-reactive protein (CRP), and advanced oxidative protein products (AOPP) as explanatory variables.

Therefore, the combination of immune responses to viral, neuronal and ZOOC antigens are strongly associated with Long COVID.

### Prediction of Comp_PP

To identify the key predictors of the Comp_PP score, we conducted three separate multiple regression analyses. The first analysis (#1) incorporated the six IgA/IgM/IgG responses to LPS and ZOOC as independent variables. Our research revealed that IgG targeting ZOOC proved to be a noteworthy factor, accounting for 13.5% of the variability in Comp_PP. **Figure 4** displays the partial regression of the Comp_PP score on IgG to ZOOC, considering age, sex, and BMI. In regression #2, Table 3 included the CRP and AOPP values. The results indicated that the combination of CRP and IgG ZOOC accounted for 36.9% of the variance in Comp_PP. Regression #3 also included the Ig responses to neuronal and viral antigens. The results show that 41.6% of the variance in Comp_PP was explained by the combined effects of CRP, IgG MPB, and IgA MBP. Therefore, once the MBP data was entered, the significance of IgG ZOOC effects diminished. It is possible that the effects of IgG on Comp_PP are influenced by autoimmunity to MBP. Therefore, we have conducted a) an analysis on the most accurate indicators of the autoimmune reactions to MBP (utilizing leaky gut and viral antigens) and ZOOC (employing the viral antigens); and b) mediation analyses whereby the effects of leaky gut on Comp_PP are mediated by autoimmunity to MBP.

**Figure 4.**
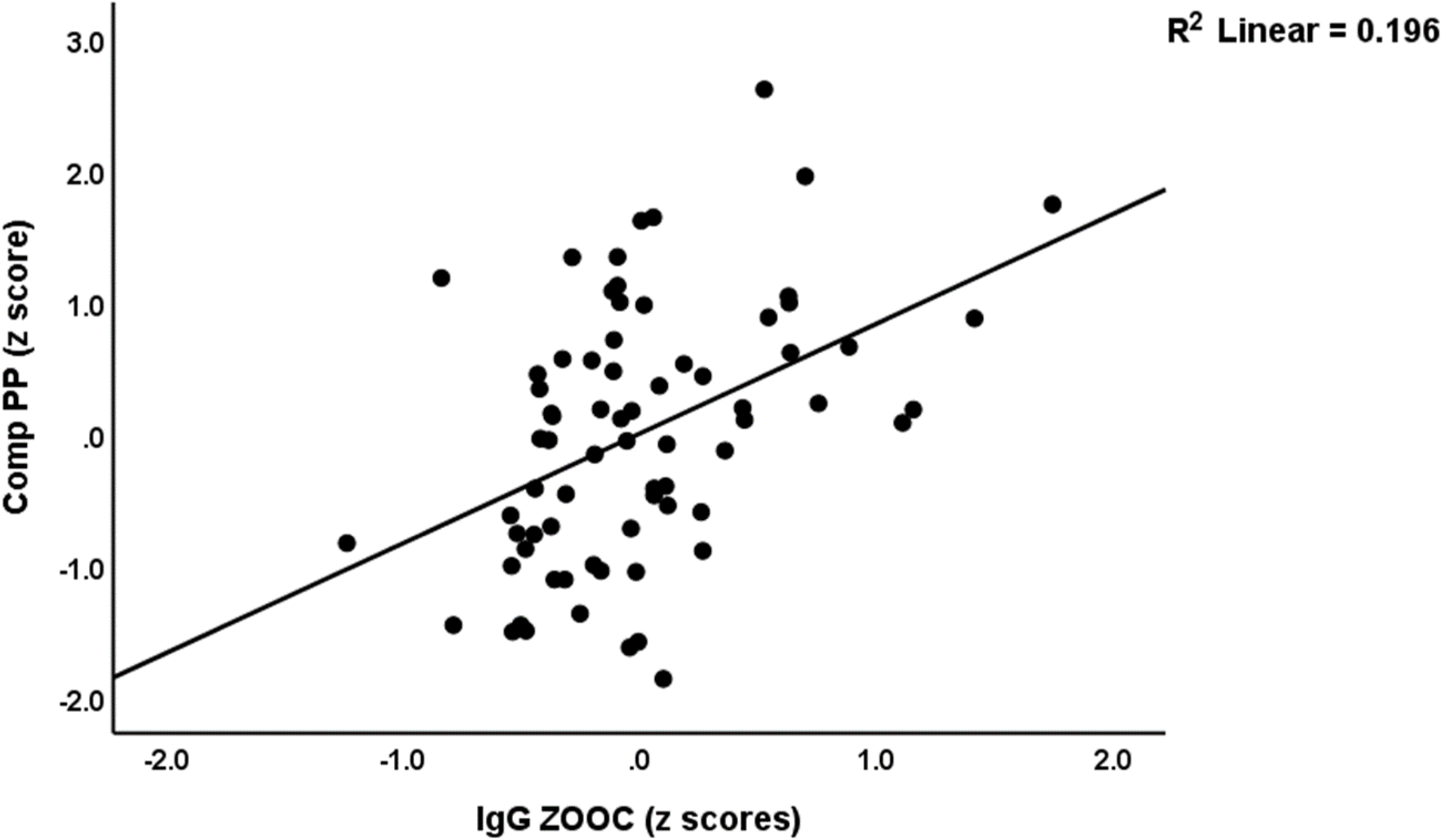
Partial regression of the physio-affective phenome of Long COVID (Comp_PP) score on IgG directed to zonulin and occludin (ZOOC); after adjustment for age, sex, and body mass index (p<0.001).

### Prediction of autoimmunity to MBP and ZOOC

We employed neural networks to accurately predict autoimmunity to MBP. The final calculation involved the combination of z-scores for IgA MBP, IgM MBP, and IgG MBP to create a composite score labeled as Comp_MBP. **Table 4**, NN#1 utilizes Comp_MBP as the output variable, with the immune responses to LPS, ZOOC, and viral antigens serving as the input variables. The model was trained using a configuration that included two hidden layers, with 5 and 4 units, respectively. The hidden layer in this model utilized the hyperbolic tangent activation function, while the output layer used the identity function. The error was greatly reduced through training. The error terms remained consistent across the training, testing, and holdout sample, suggesting that the model effectively mitigated overfitting. The precision, as determined through cross-validation, was found to be 0.750. This value represents the correlation coefficient, which indicates the strength of the relationship between the predicted and observed values. The relevance chart in **Figure 5** demonstrates the model’s assignment of predictive power to IgA, IgM, and IgG ZOOC, with IgA LPS following somewhat behind. The significance of the IgM/IgG LPS and viral antigen data was considerably lower.

**Figure 5.**
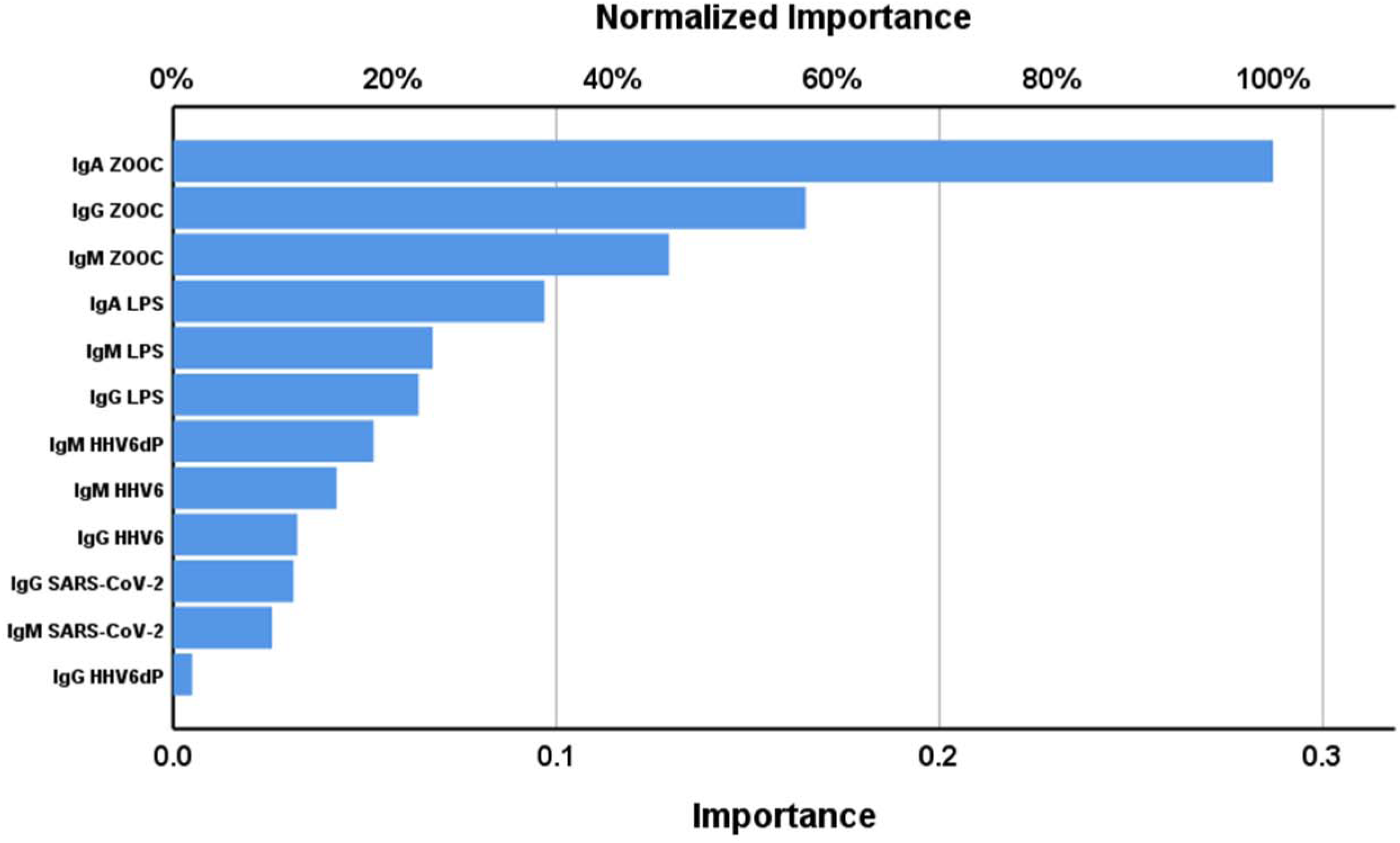
Results of neural network analysis with autoimmunity directed to myelin basic protein as output variable. Input variables are ZOOC: zonulin and occludin; LPS: lipopolysaccharides; HHV6: Human Herpes Virus 6, HHV6dP: HHV-6 duTPase; SARS-CoV-2: Severe Acute Respiratory Syndrome Coronavirus 2.

**Table 4.**
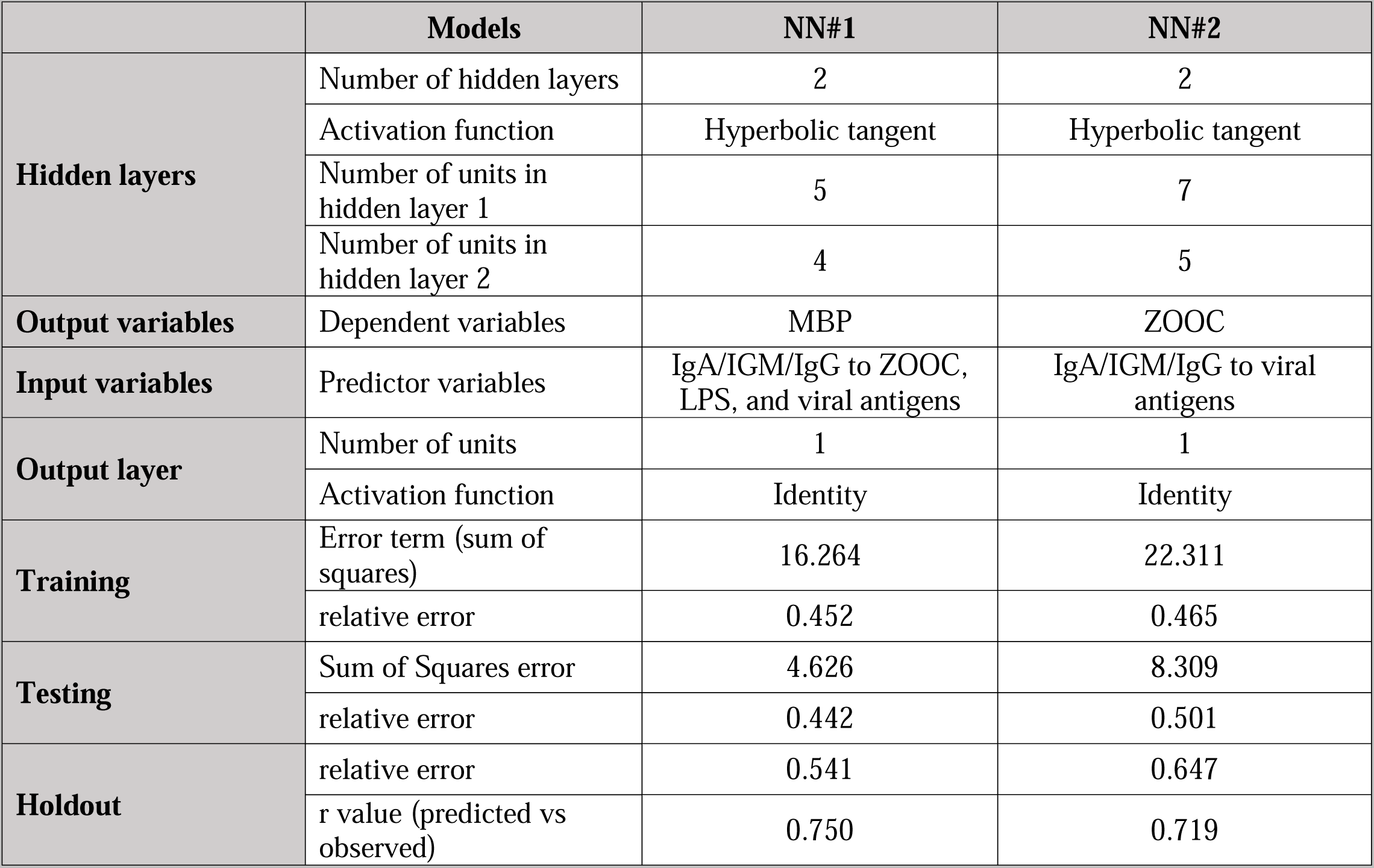
Results of neural networks (NN) with IgA/IGM/IgG directed against myelin basic protein (MBP), or zonulin and occludin (ZOOC) as dependent variables.

We have also examined the associations between the autoimmunity directed to ZOOC and BBB (both conceptualized as z unit composites based on IgA + IgG + IgM values). There was a significant correlation between both ZOOC and BBB responses (r=0.765, p<0.001, n=180). There were also significant correlations between all IgA/IgM/IgG responses to ZOOC and BBB (all at p<0.05) except between IgA-ZOOC and IgG-BBB (p=0.07, p=180).

Table 4, NN#2 uses Comp_ZOOC as output. This output is calculated by adding up the sums of the z scores of the IgA, IgM, and IgG scores, resulting in a z unit-based composite score. The input variables consisted of various immune responses to all viral antigens. Table 4, NN#2 displays comprehensive network information, including summary data and confusion matrices. The model’s precision, as determined through cross validation, was 0.719 (correlation coefficient between predicted versus observed values). Based on **Figure 6**, it can be observed that the autoimmunity targeting ZOOC is most accurately predicted by the IgA/IgM/IgG reactions to HHV-6. On the other hand, the immune responses to HHV-6 dUTPase and SARS-CoV-2 have significantly lesser significance in this context.

**Figure 6.**
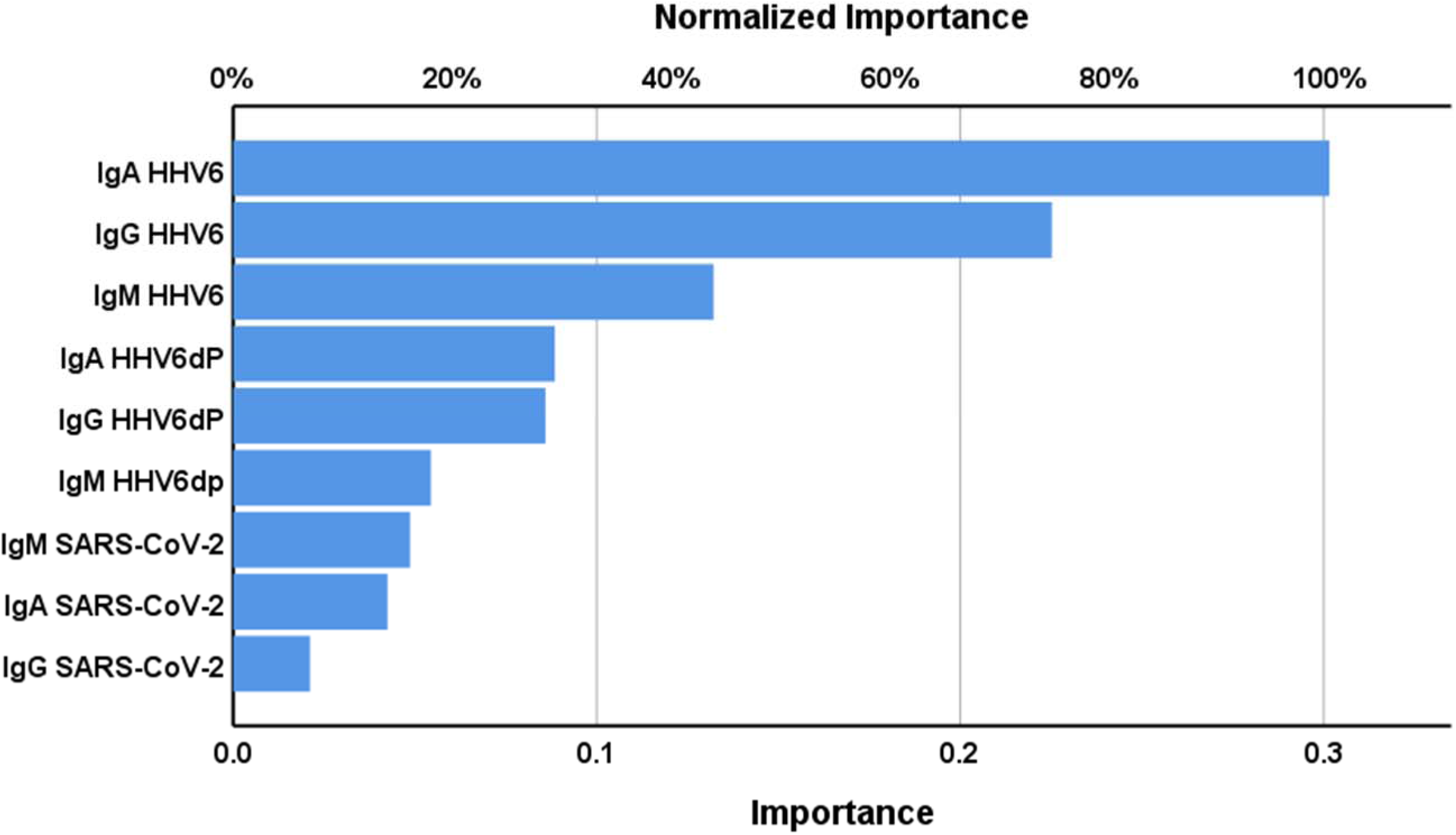
Results of neural network analysis with autoimmunity directed to zonulin and occludin as output variable. Input variables are LPS: lipopolysaccharides; HHV6: Human Herpes Virus 6, HHV6dP: HHV-6 duTPase; SARS-CoV-2: and Severe Acute Respiratory Syndrome Coronavirus 2.

**Figure 7.**
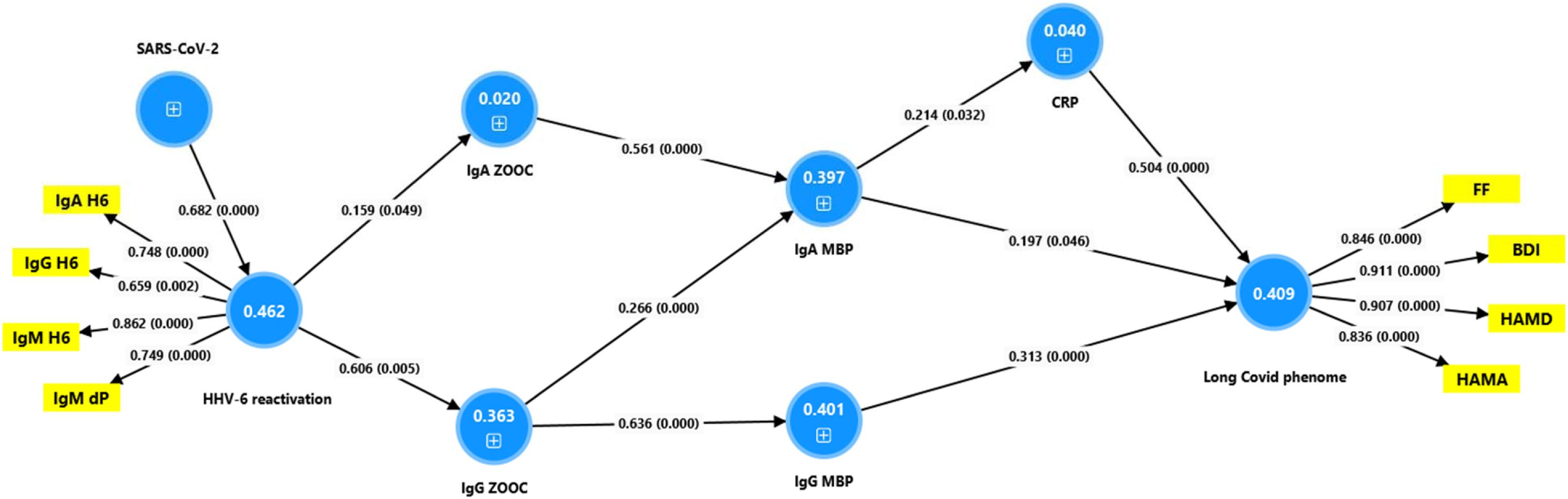
Results of Partial Least Squares analysis with the physio-affective phenome of Long COVID as final outcome variables. Input variables are C-reactive protein (CRP) levels, and autoimmunity to myelin basic protein (MBP); zonulin + occludin (ZOOC). HHV6: Human Herpes Virus 6, HHV6dP: HHV-6 duTPase; and SARS-CoV-2: Severe Acute Respiratory Syndrome Coronavirus 2. A multiple mediating model is constructed whereby HHV6 reactivation and autoimmunity to ZOOC and MBP may mediate the effects of SARS-CoV-2 infection. Shown are the explained variances (figures in blue circles), and path coefficients with exact p-value. The model shows only the significant pathways.

### Results of PLS analysis

We have conducted PLS analysis to explore the potential causal pathways between SARS-CoV-2 infection and the development of Long COVID. We employed a theoretical framework rooted in the theory outlined in our Introduction, and the results shown in Tables 3 and 4. This theory suggests that the SARS-CoV-2 infection could potentially reactivate HHV-6, leading to a condition known as leaky gut. This, in turn, may trigger autoimmune responses to neuronal antigens and inflammation. Collectively, these factors may be linked to the phenome. As a result, the PLS model produced the Long COVID phenome, which was derived as a factor extracted from the FF, BDI, HAMD, and HAMA scores. The phenome was predicted based on the analysis of immune responses to neuronal antigens and CRP levels. In addition, the immune responses to ZOOC and viral antigens were used to predict the neuronal data and CRP, while the HHV-6 assessments were used to predict the autoimmune responses to ZOOC. A latent variable called HHV-6 reactivation was created by extracting a factor from the IgA/IgM/IgG to HHV-6 and IgM to HHV-6 dUTPase. It is worth noting that the IgA/IgG responses to HHV-6 dUTPase did not have a significant impact on this factor. The data quality of this model was more than sufficient. a) The model fit indicated an SRMR of 0.059; b) the factors demonstrated Cronbach’s alpha values of 0.749 (HHV6) and 0.898 (phenome), with composite reliability values of 0.842 and 0.929, respectively; c) CTA analysis confirmed that both factors were accurately specified as reflective models; and d) The HTMT ratios indicated the presence of discriminant validity (all HTMT values were < 0.735). Our analysis revealed that CRP and IgA/IgG directed at MBP accounted for 40.9% of the variance in the Long COVID phenome. A significant portion of the data in these MBP findings was elucidated through regression analysis on the levels of IgA and/or IgG to ZOOC. The latter were predicted by the HHV-6 factor, which was found to be associated with IgM SARS-CoV-2. As such, we constructed a complex multi-step mediation model. Significant predictors of the phenome were found in the analyses of total effects. Specifically, SARS-CoV-2 (t=2.46, p=0.007), HHV-6 factor (t=2.42, p=0.008), IgG to ZOOC (t=4.36, p<0.001), and IgA to ZOOC (t=2.41, p=0.008) demonstrated noteworthy associations. The impact of SARS-CoV-2 was partially influenced by the pathway from HHV-6 to IgG ZOOC to IgG MBP, ultimately affecting the phenome (t=2.00, p=0.023). The impact of HHV-6 on the phenome was observed through the pathway from IgG ZOOC to IgG MBP, resulting in significant effects (t=1.98, p=0.024). The effects of IgG ZOOC were observed through the pathway from IgG MBP to the phenome (t=3.41 p<0.001), and the pathway from IgG ZOOC to IgA MBP to CRP to the phenome (t=1.66, p=0.049). The effects of IgA ZOOC on the phenome were found to be mediated by the path from IgA MBP to CRP, with a statistically significant t-value of 1.86 and a p-value of 0.031.

## Discussion

### The paracellular pathway and Long COVID

The first major finding of this study is that Long COVID and its physio-affective phenome exhibit dysfunctions in the paracellular pathway. This is evident through the heightened IgA/IgM/IgG-mediated autoimmune responses to zonulin and occludin. Furthermore, the analysis conducted using neural networks revealed that the levels of IgA targeting LPS combined with IgG targeting zonulin + occludin exhibited the highest predictive accuracy for Long COVID. It can be inferred that the development of leaky gut and bacterial translocation are factors contributing to Long COVID and its physio-affective symptoms.

In previous studies, it was noted that MDD and CFS both exhibit indications of heightened leaky gut and bacterial translocation, as determined by levels of IgA/IgM antibodies targeting gut commensal Gram-negative bacteria [23, 24]. Research conducted by Madison et al. [40] found that women who have successfully overcome breast cancer displayed higher levels of depression, in association with an increase in indicators of inflammation and gut permeability. The latter was determined through the use of lipopolysaccharide binding protein (LBP) as an assessment tool. Patients with inflammatory bowel disease often experience depression, which is indicated by various markers of leaky gut such as calprotectin and LBP [41]. There have been studies conducted on MDD and CFS that have found evidence of gut dysbiosis endotypes, which suggest an association with increased gut permeability and pro-inflammatory potential [42–46].

It is worth mentioning that in hospitalized COVID-19 patients, there was a correlation between elevated zonulin levels and signs of IRS activation, as well as a poorer outcome, including mortality [47]. Research conducted by Ghoshal and Ghoshal [48] suggest that individuals who have been affected by Long COVID have a higher likelihood of developing irritable bowel syndrome when compared to normal volunteers. A study utilizing Mendelian randomization demonstrated that various taxa of gut microbiota may potentially play a causal role in the development of Long COVID [49]. A separate study conducted by Su et al. [50] revealed the presence of distinct gut-microbiome enterotypes in individuals with Long COVID and its associated symptoms. In a study conducted by Mussabay et al. [51], it was found that individuals with Long COVID exhibited a notable change in their microbiome composition. Specifically, there was an observed shift towards a pro-inflammatory profile, characterized by an increased abundance of *Bacteroides*, *Faecalibacterium*, and *Prevotella*.

### Gut tight junctions and the BBB

The second major finding is that, in our study, a noteworthy correlation was observed between the autoimmune responses to tight junctions (zonulin and occludin) and the BBB (claudin and S100B). There is evidence suggesting potential damage to the BBB in COVID-19, as indicated by studies [52] and [53]. Higher levels of S100B in COVID-19 patients have been linked to the severity of their illness, indicating a potential increase in BBB permeability and potential brain damage [54]. It is worth noting that Long COVID patients may exhibit elevated levels of autoantibodies to claudin [55] and that the tight junctions of the endothelial cells of the BBB effectively prevent blood-borne factors from entering the brain [56]. Occludin and claudins play a crucial role in the tight junctions of the BBB. Interestingly, there are certain similarities in the regulation of tight junctions between the gut and the BBB [31, 56]. Furthermore, preclinical studies and studies conducted in laboratory settings indicate that zonulin may have the potential to influence the BBB [56]. However, it is important to note that this effect may not be observable in humans [57].

### HHV-6 reactivation and autoimmunity to gut tight junctions

The third major finding of this study is the strong association between HHV-6 reactivation (measured by HHV-6 and HHV-6 dUTPase IgA/IgM/IgG levels) and biomarkers indicating abnormalities in the gut tight junction. Previous studies have indicated that HHV-6 can have an impact on the gut mucosa. As an illustration, in the case of inflammatory bowel disease, HHV-6 is commonly found in inflamed mucosa and is linked to the histological disease activity and endoscopic severity of IBD [58–60]. In patients experiencing severe diarrhea after stem cell transplantation, it has been observed that HHV-6B DNA can be found in both peripheral blood mononuclear cells and large intestine tissues [59]. This suggests that HHV-6 may reactivate and infect goblet cells and histiocytes, as discovered by Amo et al. [34]. Cells positive for HHV-6 can be identified in the gastroduodenal mucosa of both immunocompetent dyspeptic patients and liver transplant recipients, as demonstrated by Halme et al. [33].

### From aberrations in tight junction to autoimmunity against neuronal antigens

Another important discovery from this study is the strong link between abnormalities in the tight junctions and increased levels of IgG/IgA/IgM-mediated autoimmunity towards neuronal antigens. This, in turn, has a significant impact on the physio-affective phenome. In a previous study, it was demonstrated that patients with MDD exhibit elevated levels of serum IgA to Gram-negative bacteria in association with increased IgG reactivity to oxidized low density lipoprotein cholesterol [28]. Furthermore, in CFS, there have been notable findings regarding the correlation between heightened indicators of bacterial translocation and elevated levels of serum IgA directed towards serotonin [61]. In both disorders, there was an observed increase in bacterial translocation, which was found to be linked to various inflammatory biomarkers. These biomarkers included lysozyme, interleukin-1β, and tumor necrosis factor [25, 28, 61]. Additionally, there was an association between increased bacterial translocation and increases in IgM responses to oxidatively modified epitopes and nitroso-adducts [28, 62]. Overall, there seem to be significant connections between gut barrier function and immune responses to neuronal and other antigens in Long COVID, its physio-affective phenome, MDD, and CFS [this study,[28, 61]].

In summary, the presence of a leaky gut and bacterial translocation could potentially play a significant role in the development of autoimmunity caused by COVID-19 and Long COVID. It has been observed that dysfunctions in the tight junctions can lead to increased bacterial translocation, which has been linked to the development of autoimmune responses and autoimmune disease [63]. An increased movement of luminal contents, such as microbiota, into the blood plays a key role in the development or worsening of human autoimmune disorders [24, 31, 63]. Especially, the translocation into the blood of gut microbes with components that have structural relatedness with the tissues or cells of the human host are involved in this process, labeled molecular mimicry [24, 31, 63]. The factors that can lead to the production of cross-reactive antibodies include various components such as LPS from Gram-negative enterobacteria, bacterial cytolethal distending toxin (CDT), Gram-positive bacteria, and various viruses [64–67]. There are distinct mechanisms by which pathogens can trigger the development of autoimmune processes. These include molecular mimicry, the exposure of cryptic antigens, superantigens, and bystander activation [68, 69]. Therefore, it may be concluded that leaky gut and resulting autoimmunity play a role in the pathophysiology of Long COVID, particularly in its physio-affective phenome. This is connected to various pathways such as IRS activation, NLRP3 inflammasome activation, oxidative stress, nitrosative stress, and more (see Introduction).

## Conclusions

This study shows that increased autoimmunity against components of the tight junctions as well as increased IgA levels to LPS of Gram-negative bacteria play a role in Long COVID and in its physio-affective phenome. Moreover, the responsivity to the tight junctions is associated with autoimmunity to neuronal antigens and the BBB. It may be suggested that dysfunctions in gut tight junctions caused by viral persistence or reactivation can lead to an increased entry of gut microbes into the bloodstream. These microbes, through structural similarities and molecular mimicry, or other mechanisms, can trigger cross-reactivity and subsequent autoimmune responses against the body’s self-epitopes or modified neo-epitopes. These processes also lead to the activation of the IRS, inflammation, oxidative stress, nitrosative stress, and increased autoimmune responses to inflammation- and oxidatively modified neo-epitopes. All those factors together determine the physio-affective phenome of Long-COVID.

## Data Availability

After receiving a suitable request and once the author has extensively analyzed the data, the lead author (MM) is willing to grant access to the SPSS file linked to this study.

## Acknowledgments

The authors are grateful to the contributors from all institutions and hospitals mentioned in this study for their support in the data collection process.

## Ethical approval and consent to participate

The Ethics Committee of the College of Medical Technology at the Islamic University of Najaf, Iraq, granted approval for this investigation (Document No. 34/2023). All procedures were consistent with Iraqi and international standards. Written informed assent was obtained from both patients and controls.

## Author’s contributions

AA and AV were responsible for blood sample collection and other patient-related tasks. Biomarker quantification in the serum was performed by AV and AFA. MM handled the statistical evaluation of the study. The manuscript was composed by MM and refined by AFA, AV, and XT with all authors reviewing and endorsing the last version.

## References

[1] Davis HE, McCorkell L, Vogel JM, Topol EJ. Long COVID: major findings, mechanisms and recommendations. Nat Rev Microbiol. 2023;21(3):133–46. 10.1038/s41579-022-00846-2.

[2] Premraj L, Kannapadi NV, Briggs J, Seal SM, Battaglini D, Fanning J, et al. Mid and long-term neurological and neuropsychiatric manifestations of post-COVID-19 syndrome: A meta-analysis. J Neurol Sci. 2022;434:120162. 10.1016/j.jns.2022.120162.

[3] Lopez-Leon S, Wegman-Ostrosky T, Perelman C, Sepulveda R, Rebolledo PA, Cuapio A, et al. More than 50 long-term effects of COVID-19: a systematic review and meta-analysis. Scientific Reports. 2021;11(1):16144. 10.1038/s41598-021-95565-8.

[4] Groff D, Sun A, Ssentongo AE, Ba DM, Parsons N, Poudel GR, et al. Short-term and Long-term Rates of Postacute Sequelae of SARS-CoV-2 Infection: A Systematic Review. JAMA Netw Open. 2021;4(10):e2128568. 10.1001/jamanetworkopen.2021.28568.

[5] Stefanou M-I, Palaiodimou L, Bakola E, Smyrnis N, Papadopoulou M, Paraskevas GP, et al. Neurological manifestations of long-COVID syndrome: a narrative review. Therapeutic Advances in Chronic Disease. 2022;13:20406223221076890. 10.1177/20406223221076890.

[6] Al-Hakeim HK, Al-Rubaye HT, Almulla AF, Al-Hadrawi DS, Maes M. Chronic Fatigue, Depression and Anxiety Symptoms in Long COVID Are Strongly Predicted by Neuroimmune and Neuro-Oxidative Pathways Which Are Caused by the Inflammation during Acute Infection. In: Journal of Clinical Medicine. 12. 2023.

[7] Al-Hakeim HK, Al-Rubaye HT, Al-Hadrawi DS, Almulla AF, Maes M. Long-COVID post-viral chronic fatigue and affective symptoms are associated with oxidative damage, lowered antioxidant defenses and inflammation: a proof of concept and mechanism study. Molecular Psychiatry. 2022. 10.1038/s41380-022-01836-9.

[8] Maes M, Al-Rubaye HT, Almulla AF, Al-Hadrawi DS, Stoyanova K, Kubera M, et al. Lowered Quality of Life in Long COVID Is Predicted by Affective Symptoms, Chronic Fatigue Syndrome, Inflammation and Neuroimmunotoxic Pathways. In: International Journal of Environmental Research and Public Health. 19. 2022.

[9] Al-Hadrawi DS, Al-Rubaye HT, Almulla AF, Al-Hakeim HK, Maes M. Lowered oxygen saturation and increased body temperature in acute COVID-19 largely predict chronic fatigue syndrome and affective symptoms due to Long COVID: A precision nomothetic approach. Acta Neuropsychiatrica. 2022:1–12. 10.1017/neu.2022.21.

[10] Al-Jassas HK, Al-Hakeim HK, Maes M. Intersections between pneumonia, lowered oxygen saturation percentage and immune activation mediate depression, anxiety, and chronic fatigue syndrome-like symptoms due to COVID-19: A nomothetic network approach. J Affect Disord. 2022;297:233–45. 10.1016/j.jad.2021.10.039.

[11] Al-Hakeim HK, Khairi Abed A, Rouf Moustafa S, Almulla AF, Maes M. Tryptophan catabolites, inflammation, and insulin resistance as determinants of chronic fatigue syndrome and affective symptoms in long COVID. Frontiers in Molecular Neuroscience. 2023;16.

[12] Al-Hakeim HK, Abed AK, Almulla AF, Rouf Moustafa S, Maes M. Anxiety due to Long COVID is partially driven by activation of the tryptophan catabolite (TRYCAT) pathway. Asian Journal of Psychiatry. 2023;88:103723. 10.1016/j.ajp.2023.103723.

[13] Abbas FA, Yanin T, Bo Z, Aristo V, Michael M. Immune activation and immune-associated neurotoxicity in Long-COVID: A systematic review and meta-analysis of 82 studies comprising 58 cytokines/chemokines/growth factors. medRxiv. 2024:2024.02.08.24302516. 10.1101/2024.02.08.24302516.

[14] Yong SJ, Halim A, Halim M, Liu S, Aljeldah M, Al Shammari BR, et al. Inflammatory and vascular biomarkers in post-COVID-19 syndrome: A systematic review and meta-analysis of over 20 biomarkers. Reviews in Medical Virology. 2023;33(2):e2424. 10.1002/rmv.2424.

[15] Vojdani A, Almulla AF, Zhou B, Al-Hakeim HK, Maes M. Reactivation of herpesvirus type 6 and IgA/IgM-mediated responses to activin-A underpin long COVID, including affective symptoms and chronic fatigue syndrome. Acta Neuropsychiatr. 2024:1–13. 10.1017/neu.2024.10.

[16] Vojdani A, Vojdani E, Saidara E, Maes M. Persistent SARS-CoV-2 Infection, EBV, HHV-6 and Other Factors May Contribute to Inflammation and Autoimmunity in Long COVID. In: Viruses. 15. 2023.

[17] Su Y, Yuan D, Chen DG, Ng RH, Wang K, Choi J, et al. Multiple early factors anticipate post-acute COVID-19 sequelae. Cell. 2022;185(5):881–95.e20. 10.1016/j.cell.2022.01.014.

[18] Almulla AF, Maes M, Bo Z, Hussein KA-H, Aristo V. Brain-targeted autoimmunity is strongly associated with Long COVID and its chronic fatigue syndrome as well as its affective symptoms. medRxiv. 2023:2023.10.04.23296554. 10.1101/2023.10.04.23296554.

[19] Morris G, Maes M. Oxidative and Nitrosative Stress and Immune-Inflammatory Pathways in Patients with Myalgic Encephalomyelitis (ME)/Chronic Fatigue Syndrome (CFS). Curr Neuropharmacol. 2014;12(2):168–85. 10.2174/1570159X11666131120224653.

[20] Morris G, Maes M. Myalgic encephalomyelitis/chronic fatigue syndrome and encephalomyelitis disseminata/multiple sclerosis show remarkable levels of similarity in phenomenology and neuroimmune characteristics. BMC Medicine. 2013;11(1):205. 10.1186/1741-7015-11-205.

[21] Maes M, Carvalho AF. The Compensatory Immune-Regulatory Reflex System (CIRS) in Depression and Bipolar Disorder. Mol Neurobiol. 2018;55(12):8885–903. 10.1007/s12035-018-1016-x.

[22] Almulla AF, Abbas Abo Algon A, Tunvirachaisakul C, Al-Hakeim HK, Maes M. T helper-1 activation via interleukin-16 is a key phenomenon in the acute phase of severe, first-episode major depressive disorder and suicidal behaviors. J Adv Res. 2023. 10.1016/j.jare.2023.11.012.

[23] Maes M, Mihaylova I, Leunis JC. Increased serum IgA and IgM against LPS of enterobacteria in chronic fatigue syndrome (CFS): indication for the involvement of gram-negative enterobacteria in the etiology of CFS and for the presence of an increased gut-intestinal permeability. J Affect Disord. 2007;99(1-3):237–40. 10.1016/j.jad.2006.08.021.

[24] Maes M, Kubera M, Leunis JC. The gut-brain barrier in major depression: intestinal mucosal dysfunction with an increased translocation of LPS from gram negative enterobacteria (leaky gut) plays a role in the inflammatory pathophysiology of depression. Neuro Endocrinol Lett. 2008;29(1):117–24.

[25] Simeonova D, Stoyanov D, Leunis JC, Carvalho AF, Kubera M, Murdjeva M, et al. Increased Serum Immunoglobulin Responses to Gut Commensal Gram-Negative Bacteria in Unipolar Major Depression and Bipolar Disorder Type 1, Especially When Melancholia Is Present. Neurotox Res. 2020;37(2):338–48. 10.1007/s12640-019-00126-7.

[26] Simeonova D, Ivanovska M, Murdjeva M, Carvalho AF, Maes M. Recognizing the Leaky Gut as a Trans-diagnostic Target for Neuroimmune Disorders Using Clinical Chemistry and Molecular Immunology Assays. Curr Top Med Chem. 2018;18(19):1641–55. 10.2174/1568026618666181115100610.

[27] Ohlsson L, Gustafsson A, Lavant E, Suneson K, Brundin L, Westrin Å, et al. Leaky gut biomarkers in depression and suicidal behavior. Acta Psychiatr Scand. 2019;139(2):185–93. 10.1111/acps.12978.

[28] Maes M, Kubera M, Leunis JC, Berk M, Geffard M, Bosmans E. In depression, bacterial translocation may drive inflammatory responses, oxidative and nitrosative stress (O&NS), and autoimmune responses directed against O&NS-damaged neoepitopes. Acta Psychiatrica Scandinavica. 2013;127(5):344–54. 10.1111/j.1600-0447.2012.01908.x.

[29] Rudzki L, Maes M. The Microbiota-Gut-Immune-Glia (MGIG) Axis in Major Depression. Mol Neurobiol. 2020;57(10):4269–95. 10.1007/s12035-020-01961-y.

[30] Vojdani A, Vojdani E, Kharrazian D. Fluctuation of zonulin levels in blood vs stability of antibodies. World J Gastroenterol. 2017;23(31):5669–79. 10.3748/wjg.v23.i31.5669.

[31] Vojdani A, Vojdani E. Food-associated autoimmunities: when food breaks your immune system. A&G Press; 2019.

[32] Fasano A. Intestinal permeability and its regulation by zonulin: diagnostic and therapeutic implications. Clin Gastroenterol Hepatol. 2012;10(10):1096–100. 10.1016/j.cgh.2012.08.012.

[33] Halme L, Arola J, Höckerstedt K, Lautenschlager I. Human Herpesvirus 6 Infection of the Gastroduodenal Mucosa. Clinical Infectious Diseases. 2008;46(3):434–9. 10.1086/525264.

[34] Amo K, Tanaka-Taya K, Inagi R, Miyagawa H, Miyoshi H, Okusu I, et al. Human herpesvirus 6B infection of the large intestine of patients with diarrhea. Clin Infect Dis. 2003;36(1):120–3. 10.1086/345464.

[35] World Health Organization W. A clinical case definition of post COVID-19 condition by a Delphi consensus 6 October. WHO. 2021.

[36] Hamilton M. The assessment of anxiety states by rating. Br J Med Psychol. 1959;32(1):50–5. 10.1111/j.2044-8341.1959.tb00467.x.

[37] Hamilton M. A rating scale for depression. J Neurol Neurosurg Psychiatry. 1960;23(1):56–62. 10.1136/jnnp.23.1.56.

[38] Hautzinger MKFKhCBATSRABGK. Beck Depressions-Inventar: BDI-II; Revision; Manual. Frankfurt am Main: Pearson; 2009.

[39] Zachrisson O, Regland B, Jahreskog M, Kron M, Gottfries CG. A rating scale for fibromyalgia and chronic fatigue syndrome (the FibroFatigue scale). Journal of Psychosomatic Research. 2002;52(6):501–9. 10.1016/S0022-3999(01)00315-4.

[40] Madison AA, Andridge R, Kantaras AH, Renna ME, Bennett JM, Alfano CM, et al. Depression, Inflammation, and Intestinal Permeability: Associations with Subjective and Objective Cognitive Functioning throughout Breast Cancer Survivorship. Cancers (Basel). 2023;15(17). 10.3390/cancers15174414.

[41] Iordache MM, Tocia C, Aschie M, Dumitru A, Manea M, Cozaru GC, et al. Intestinal Permeability and Depression in Patients with Inflammatory Bowel Disease. J Clin Med. 2022;11(17). 10.3390/jcm11175121.

[42] Maes M, Vasupanrajit A, Jirakran K, Klomkliew P, Chanchaem P, Tunvirachaisakul C, et al. Adverse childhood experiences and reoccurrence of illness impact the gut microbiome, which affects suicidal behaviours and the phenome of major depression: towards enterotypic phenotypes. Acta Neuropsychiatr. 2023;35(6):328–45. 10.1017/neu.2023.21.

[43] Stallmach A, Quickert S, Puta C, Reuken PA. The gastrointestinal microbiota in the development of ME/CFS: a critical view and potential perspectives. Front Immunol. 2024;15:1352744. 10.3389/fimmu.2024.1352744.

[44] Wang JH, Choi Y, Lee JS, Hwang SJ, Gu J, Son CG. Clinical evidence of the link between gut microbiome and myalgic encephalomyelitis/chronic fatigue syndrome: a retrospective review. Eur J Med Res. 2024;29(1):148. 10.1186/s40001-024-01747-1.

[45] Chen CY, Wang YF, Lei L, Zhang Y. Impacts of microbiota and its metabolites through gut-brain axis on pathophysiology of major depressive disorder. Life Sci. 2024;351:122815. 10.1016/j.lfs.2024.122815.

[46] Tao K, Yuan Y, Xie Q, Dong Z. Relationship between human oral microbiome dysbiosis and neuropsychiatric diseases: An updated overview. Behavioural Brain Research. 2024:115111.

[47] Palomino-Kobayashi LA, Ymaña B, Ruiz J, Mayanga-Herrera A, Ugarte-Gil MF, Pons MJ. Zonulin, a marker of gut permeability, is associated with mortality in a cohort of hospitalised peruvian COVID-19 patients. Front Cell Infect Microbiol. 2022;12:1000291. 10.3389/fcimb.2022.1000291.

[48] Ghoshal UC, Ghoshal U. Gastrointestinal involvement in post-acute Coronavirus disease (COVID)-19 syndrome. Curr Opin Infect Dis. 2023;36(5):366–70. 10.1097/qco.0000000000000959.

[49] Li Z, Xia Q, Feng J, Chen X, Wang Y, Ren X, et al. The causal role of gut microbiota in susceptibility of Long COVID: a Mendelian randomization study. Front Microbiol. 2024;15:1404673. 10.3389/fmicb.2024.1404673.

[50] Su Q, Lau RI, Liu Q, Li MKT, Yan Mak JW, Lu W, et al. The gut microbiome associates with phenotypic manifestations of post-acute COVID-19 syndrome. Cell Host Microbe. 2024;32(5):651–60.e4. 10.1016/j.chom.2024.04.005.

[51] Mussabay K, Kozhakhmetov S, Dusmagambetov M, Mynzhanova A, Nurgaziyev M, Jarmukhanov Z, et al. Gut Microbiome and Cytokine Profiles in Post-COVID Syndrome. Viruses. 2024;16(5). 10.3390/v16050722.

[52] Hernández-Parra H, Reyes-Hernández OD, Figueroa-González G, González-Del Carmen M, González-Torres M, Peña-Corona SI, et al. Alteration of the blood-brain barrier by COVID-19 and its implication in the permeation of drugs into the brain. Front Cell Neurosci. 2023;17.

[53] Krasemann S, Haferkamp U, Pfefferle S, Woo MS, Heinrich F, Schweizer M, et al. The blood-brain barrier is dysregulated in COVID-19 and serves as a CNS entry route for SARS-CoV-2. Stem Cell Reports. 2022;17(2):307–20. 10.1016/j.stemcr.2021.12.011.

[54] Aceti A, Margarucci LM, Scaramucci E, Orsini M, Salerno G, Di Sante G, et al. Serum S100B protein as a marker of severity in Covid-19 patients. Scientific Reports. 2020;10(1):18665. 10.1038/s41598-020-75618-0.

[55] Fonseca DLM, Filgueiras IS, Marques AHC, Vojdani E, Halpert G, Ostrinski Y, et al. Severe COVID-19 patients exhibit elevated levels of autoantibodies targeting cardiolipin and platelet glycoprotein with age: a systems biology approach. npj Aging. 2023;9(1):21. 10.1038/s41514-023-00118-0.

[56] Liu WY, Wang ZB, Zhang LC, Wei X, Li L. Tight junction in blood-brain barrier: an overview of structure, regulation, and regulator substances. CNS Neurosci Ther. 2012;18(8):609–15. 10.1111/j.1755-5949.2012.00340.x.

[57] Stuart CM, Varatharaj A, Winberg ME, Galea P, Larsson HBW, Cramer SP, et al. Zonulin and blood-brain barrier permeability are dissociated in humans. Clin Transl Med. 2022;12(7):e965. 10.1002/ctm2.965.

[58] Nahar S, Hokama A, Fujita J. Clinical significance of cytomegalovirus and other herpes virus infections in ulcerative colitis. Pol Arch Intern Med. 2019;129(9):620–6. 10.20452/pamw.14835.

[59] Nahar S, Iraha A, Hokama A, Uehara A, Parrott G, Ohira T, et al. Evaluation of a multiplex PCR assay for detection of cytomegalovirus in stool samples from patients with ulcerative colitis. World J Gastroenterol. 2015;21(44):12667–75. 10.3748/wjg.v21.i44.12667.

[60] Mousset S, Martin H, Berger A, Heß S, Bug G, Kriener S, et al. Human herpesvirus 6 in biopsies from patients with gastrointestinal symptoms after allogeneic stem cell transplantation. Ann Hematol. 2012;91(5):737–42. 10.1007/s00277-011-1354-5.

[61] Maes M, Ringel K, Kubera M, Anderson G, Morris G, Galecki P, et al. In myalgic encephalomyelitis/chronic fatigue syndrome, increased autoimmune activity against 5-HT is associated with immuno-inflammatory pathways and bacterial translocation. Journal of Affective Disorders. 2013;150(2):223–30. 10.1016/j.jad.2013.03.029.

[62] Maes M, Simeonova D, Stoyanov D, Leunis JC. Upregulation of the nitrosylome in bipolar disorder type 1 (BP1) and major depression, but not BP2: Increased IgM antibodies to nitrosylated conjugates are associated with indicants of leaky gut. Nitric Oxide. 2019;91:67–76. 10.1016/j.niox.2019.07.003.

[63] English J, Connolly L, Stewart LD. Increased Intestinal Permeability: An Avenue for the Development of Autoimmune Disease? Exposure and Health. 2024;16(2):575–605. 10.1007/s12403-023-00578-5.

[64] Thisayakorn P, Thipakorn Y, Tantavisut S, Sirivichayakul S, Vojdani A, Maes M. Increased IgA-mediated responses to the gut paracellular pathway and blood-brain barrier proteins predict delirium due to hip fracture in older adults. Front Neurol. 2024;15:1294689. 10.3389/fneur.2024.1294689.

[65] Cusick MF, Libbey JE, Fujinami RS. Molecular mimicry as a mechanism of autoimmune disease. Clin Rev Allergy Immunol. 2012;42(1):102–11. 10.1007/s12016-011-8294-7.

[66] Oldstone MB. Molecular mimicry and autoimmune disease. Cell. 1987;50(6):819-20. 10.1016/0092-8674(87)90507-1.

[67] McRae BL, Vanderlugt CL, Dal Canto MC, Miller SD. Functional evidence for epitope spreading in the relapsing pathology of experimental autoimmune encephalomyelitis. J Exp Med. 1995;182(1):75–85. 10.1084/jem.182.1.75.

[68] Fujinami RS, Oldstone MB, Wroblewska Z, Frankel ME, Koprowski H. Molecular mimicry in virus infection: crossreaction of measles virus phosphoprotein or of herpes simplex virus protein with human intermediate filaments. Proc Natl Acad Sci U S A. 1983;80(8):2346–50. 10.1073/pnas.80.8.2346.

[69] Scherer MT, Ignatowicz L, Winslow GM, Kappler JW, Marrack P. Superantigens: bacterial and viral proteins that manipulate the immune system. Annu Rev Cell Biol. 1993;9:101–28. 10.1146/annurev.cb.09.110193.000533.

